# The Validity and Reliability of Dichotomized Self-rated Health Under Different Cutpoints

**DOI:** 10.1101/2024.04.18.24306035

**Authors:** Charles Plante, Sharalynn Missiuna, Cordell Neudorf

**Affiliations:** Research Department, Saskatchewan Health Authority, Saskatoon, Canada; Department of Community Health and Epidemiology, University of Saskatchewan, Saskatoon, Canada; College of Medicine, University of Saskatchewan, Saskatoon, Canada

**Keywords:** Health Surveys, Epidemiology, Reliability And Validity, Data Analysis, Health Status Indicators

## Abstract

Self-rated health is a widely used indicator of overall health status. It is most often reported on a Likert scale of three to five values in surveys. To facilitate presentation and interpretation, it is common practice to simplify the variable by dichotomizing it; however, there has been little documented reflection on how this should be done. This paper explores all four possible dichotomizations of self-reported health, taken from three years of the Canadian Community Health Survey and reported by a Likert scale. We evaluated each dichotomization stratified by sociodemographic variables. We use regression analysis to explore the validity and reliability of all four possible dichotomizations by mapping them to the Health Utility Index. We found that lower cutpoints of dichotomization capture more pronounced differences in health status and are more consistent across sociodemographic variables. However, higher cutpoints of dichotomization should be considered for small data sets.

**About the Research Department:** The Saskatchewan Health Authority Research Department leads collaborative research to enhance Saskatchewan’s health and healthcare. We provide diverse research services to SHA staff, clinicians, and team members, including surveys, study design, database development, statistical analysis, and assistance with research funding. We also spearhead our own research programs to strengthen research and analytic capability and learning within Saskatchewan’s health system.

**About the UPHN:** The Urban Public Health Network (UPHN) is a national organization established in 2004 which today includes the Medical Officers of Health in 24 of Canada’s large urban centres. Working collaboratively and with a collective voice, the network addresses public health issues that are common to urban populations. Research operations of the UPHN are conducted in partnership with the University of Saskatchewan.

**Disclaimer:** This working paper is for discussion and comment purposes. It has not been peer-reviewed nor been subject to review by Research Department staff or executives. Any opinions expressed in this paper are those of the author(s) and not those of the Saskatchewan Health Authority.

**Suggested Citation:** Charles Plante, Sharalynn Missiuna, and Cordell Neudorf. 2024. “The Validity and Reliability of Dichotomized Self-rated Health Under Different Cutpoints.” medRxiv.

**Extended Abstract:** *Introduction:* Self-rated health is a widely used indicator of overall health status. It is most often reported on a Likert scale of three to five values in surveys. To facilitate presentation and interpretation, it is common practice to simplify the variable by dichotomizing it; however, little documented reflection has been done on how this should be done.

*Methods:* We use regression analysis to explore the validity and reliability of all four possible dichotomizations of self-reported health in the Canadian Community Health Survey in 2013-2015 by mapping them to a validated health measure: the Health Utility Index Mark 3 (HUI). We posit that more valid cutpoints in self-rated health are associated with larger changes in HUI. We posit further that more reliable cutpoints are associated with similar changes across sociodemographic variables, including age, sex, education, marital status, geography and income. We also provide descriptive statistics to contextualize our analysis.

*Results:* The greatest proportion of respondents reported having “very good” health, although the proportion of the population reporting “excellent” or “very good” health decreased with age. Similarly, Canadians tend to score highly in HUI. Our regression results suggest that HUI tends to be higher for younger, richer, married, educated and urban populations. However, these associations are muted as the cutpoint used to dichotomize self-reported health is raised. The model with the lowest cutpoint, distinguishing between poor health and all other health statuses, was associated with the greatest and most consistent negative changes in HUI among different sociodemographic groups.

*Conclusions:* Dichotomizing self-rated health using lower cutpoints captures more pronounced differences in health status measured by HUI and tends to capture more consistent differences across sociodemographic variables. That is, lower cutpoints produce more valid and reliable results. However, lower cutpoints isolate less commonly reported health levels and may lead to less accurate results in smaller populations.

**Key Points:**

- This article addresses the knowledge gap concerning the most accurate way to dichotomize self-rated health data reported using a Likert scale.
- This paper explores the validity and reliability of all four possible dichotomizations of self-reported health reported by a Likert scale.
- Lower cutpoints of dichotomization capture more pronounced differences in health status and are more consistent across sociodemographic variables.
- Higher cutpoints of dichotomization should be considered for small data sets.

## Introduction

Self-rated health is a widely used indicator of overall health status. Several studies have validated its usefulness as a subjective proxy for objective health measures.^1–9^ In surveys, self-rated health is most often reported as a Likert scale consisting of three to five values ranging from higher values like “excellent” or “very good” to lower values like “very poor” or “very bad.” The Canadian Community Health Survey (CCHS)—a survey that is used widely in health surveillance and research in Canada—asks respondents whether “in general” they would say their health is: “excellent,” “very good,” “good,” “fair,” or “poor.” Best statistical practice dictates that when we work with self-rated health, we should use models and methods that respect the ordinality of the variable;^10^ however, when analyzing dissemination among less specialized audiences, it has become common practice to simplify the variable by dichotomizing it. For example, in their 2008 examination of health inequalities in cities, the Canadian Institute for Health Information (CIHI)^11^ dichotomized responses to the aforementioned CCHS question by grouping values above and below a cutpoint between “very good” and “good.” Despite the popularity of this kind of practice, there has been surprisingly little documented consideration of how this should be done. For instance, should CIHI have dichotomized self-rated health the way it did? Or should they have chosen a different cutpoint?

In this paper, we use the Health Utility Index Mark 3 (HUI)^12,13^ to test the validity and reliability of different dichotomizations of self-rated health in three years of the CCHS. Although we are not the first to consider how best self-rated health should be dichotomized, we approach the problem more systematically than in previous studies. For instance, to the best of our knowledge, we are the first to consider this question by examining their association with a measure of “actual”^1^ health functioning. Finnas et al.^14^ and Bourne^15^ report on how different dichotomizations of self-rated health result in different summary findings in Finland and Jamaica across various sociodemographic variables, including age, sex, marital status, education, income, and/or geography. The HUI is a comprehensive and leading measure of health-related quality of life developed based on multi-attribute utility theory.^10,12^

HUI has been validated for children and adults living with a variety of conditions. ^16–19^ Briefly, it collects information from respondents via a series of 32 questions and then uses algorithms to generate scores in eight domains of health and well-being. These scores are combined to give a summary score of the respondent’s overall health-related quality of life.^20^ Nonetheless, the HUI is not without shortcomings. Specifically, it has been criticized for focusing too narrowly on functional limitations.^21,22^ Also, for potentially overestimating changes in mental health.^21^

## Methods

### Data source

The CCHS is an annual national cross-sectional survey of approximately 65 000 Canadians at least 12 years old. The survey does not include individuals living in the territories, living in Aboriginal settlements, full-time members of the Canadian Forces, and residents of institutions.^2^ To carry out our analysis, we pooled annual components of the CCHS over 2013-2015, the most recent years we had access to in which the HUI was collected in all provinces (the CCHS routinely collects self-rated health in all provinces). We additionally restricted our sample to individuals aged 15-74 and individuals who provided complete responses to all the sociodemographic variables considered in our study. Our final study sample consisted of just over 130 000 responses. We accessed and analyzed the CCHS via Statistics Canada’s Research Data Centres program.

### Health variables

As mentioned, self-rated health is collected in the CCHS as a Likert scale, allowing the following responses: “excellent,” “very good,” “good,” “fair,” or “poor.” This variable accommodates four cutpoints and so four ways of dichotomizing self-rated health. Working with these cutpoints, we elected to operationalize four categories of lesser health: “poor health,” “poor to fair health,” “poor to good health,” and “poor to very good health.” Note that zero values for each of these variables report greater health. The HUI is scored on a continuous scale where 0 indicates death, and 1 is perfect health. Scores of less than 0 are possible and denote health considered to be worse than death. We made no modifications to HUI.

### Sociodemographic variables

In this study, we grouped and coded sociodemographic variables in the CCHS as follows: age: 15-24, 25-34, 35-44, 45-54, 55-64, and 65-74; sex self-identification: male and female; education level: “less than secondary school graduation,” “secondary school graduation,” and “more than secondary school education”; marital status: “married or common-law” and “other marital status”; population density: “urban” and “rural”; and, individual self-rated income quintile: 1-5, with the first being the poorest and the last the richest. We grouped sociodemographic variable values to reduce the complexity of the analysis and facilitate interpretation and reporting.

### Analysis

To investigate the validity and reliability of different dichotomizations of self-rated health, we estimated a) to what extent they predicted large changes in HUI, and b) whether they did so consistently across different subpopulations. We estimated four separate regressions, one for each dichotomization of self-rated health. Each model regressed HUI on the dichotomization of self-rated health, sociodemographic variables, and interactions between the dichotomization of self-rated health and sociodemographic variables. The estimated intercept in these models is the predicted level of HUI for the reference population when the dichotomization of self-rated health is set to 0 (i.e. greater health). The direct effect of each sociodemographic variable indicates the extent to which each is associated with different levels of HUI independently of how they associate their health status with different levels of self-rated health (thus pushing the intercept upward or downward). A large estimated effect of the dichotomization of self-rated health on HUI supports the validity of self-reported health. The interaction effect of each sociodemographic variable indicates the extent to which the effect of the dichotomization of self-rated health tends to vary relative to the reference group for each sociodemographic variable. Thus, large and consistent interactions suggest that the dichotomization of self-rated health does not capture the same level of change in HUI across different groups and, therefore, is less reliable. Bootstrapping was used to calculate all standard errors, and all estimates were weighted by the probability weights provided with the CCHS.

To contextualize our analysis, we also calculated descriptive statistics for each sociodemographic variable and prevalence rates of each dichotomization of self-rated health for each sociodemographic variable.

## Results

Figure 1 illustrates the distribution of self-rated health by sex and age. In all age groups, relatively few respondents reported having “poor” or “fair health,” whereas more respondents tended to report having “good,” “very good,” or “excellent” health. The greatest proportion of respondents reported having “very good” health. As age increased, the proportion of individuals reporting “poor” or “fair” health tended to increase, while the proportion of individuals reporting “very good” or “excellent” health decreased. There were not large differences in the distribution of self-rated health across sexes.

**Figure 1.**
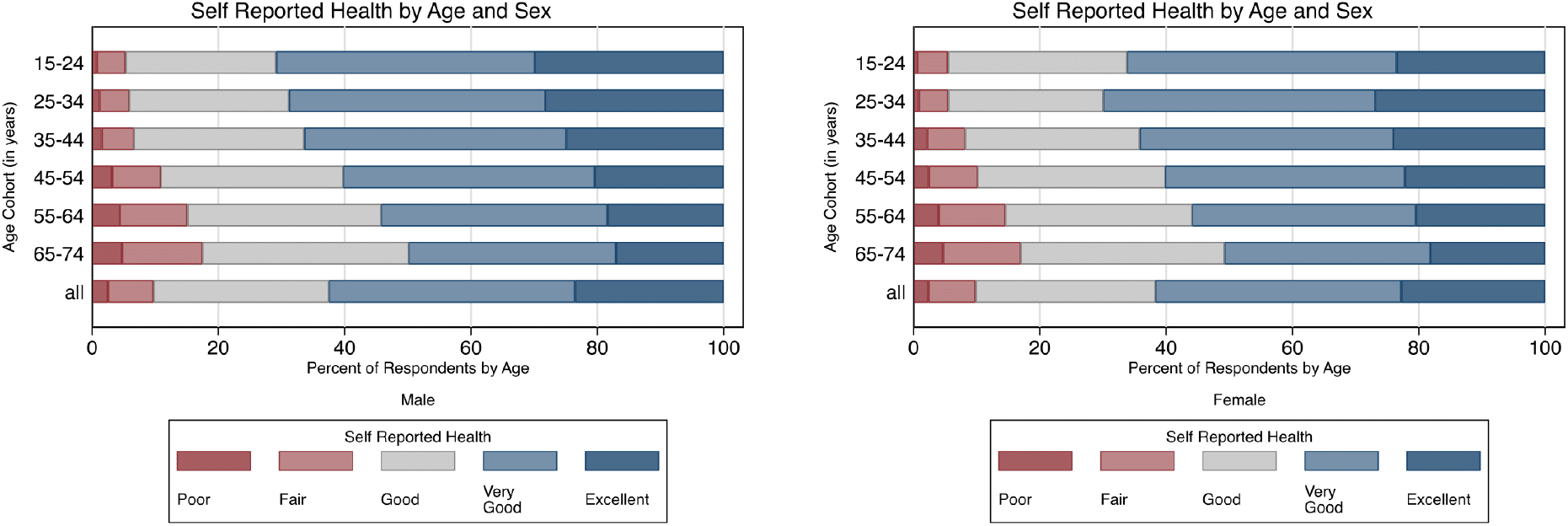
Percent self-rated health by sex and age in the pooled Canadian Community Health Survey sample, 2013-2015. Notes. Data Source: Canadian Community Health Survey 2013- 2015, Statistics Canada, and author’s calculations

The distributions of each sociodemographic variable in the pooled CCHS sample are presented in Table 1. Table 2 reports the prevalence of each dichotomization of self-rated health within each sub-population. Shading in Table 2 has been carried out independently within each dichotomization of self-rated health, and darker shading represents greater prevalences of lesser health. Across all four dichotomizations, the greatest prevalences are associated with the subpopulations with the lowest income and the lowest education, particularly within older populations. Although, overall, older subpopulations tend to have higher prevalences, these are less pronounced when income and education are considered. Absolute differences in these prevalences tend to be large for dichotomizations of self-rated health that use greater cutpoints.

**Table 1.**
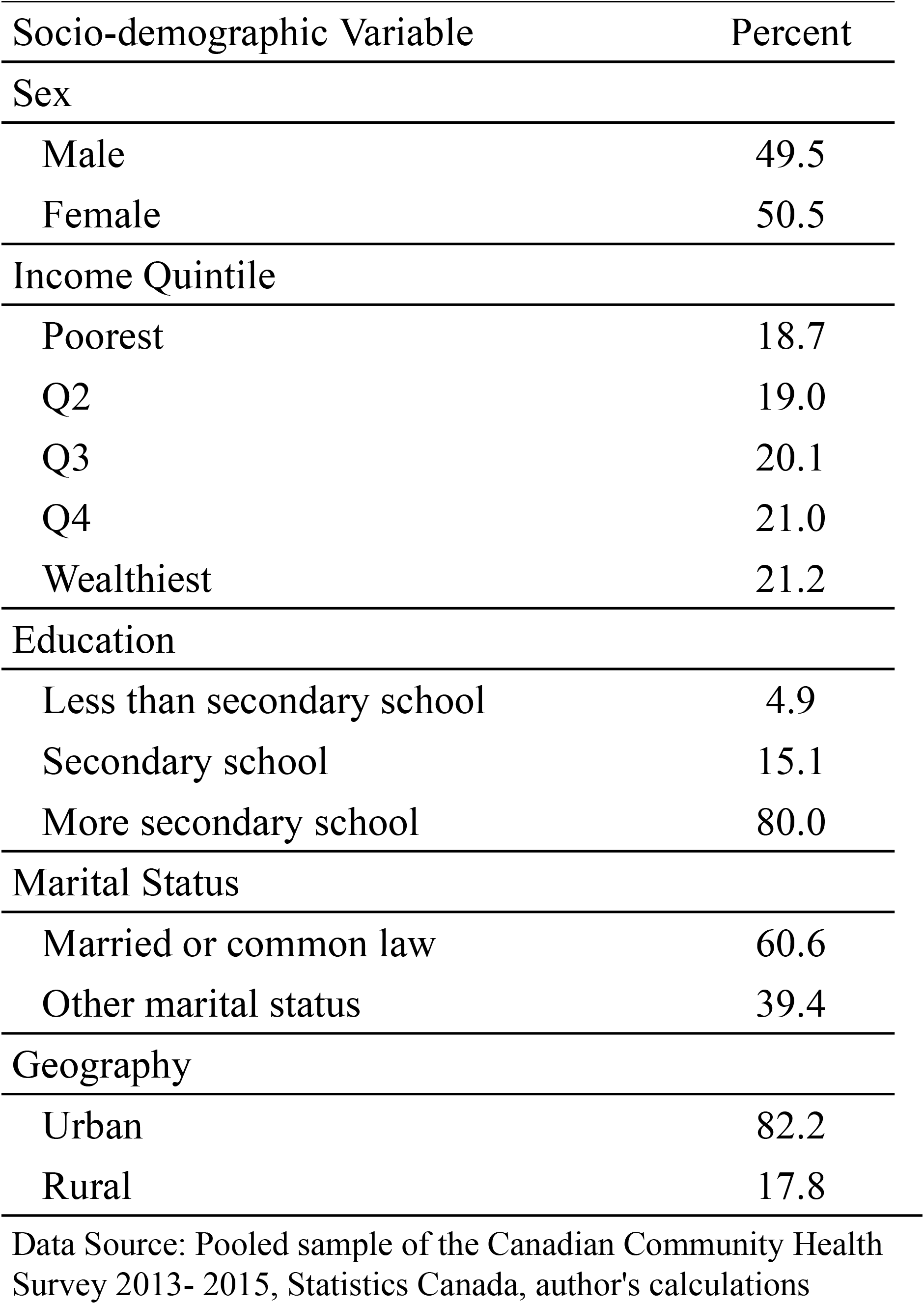
Weighted distribution of socio-demographic variables in Canadian Community Health Survey sample, 2013-2015.

**Table 2.** Percent of different sub-populations reporting lesser health under different dichotomizations of self-rated health in Canadian Community Health Survey, 2013-2015.

|  | Percent of Population Reporting Poor Health |  |  |  |  |  |  | Percent of Population Reporting Poor Health or Fair Health |  |  |  |  |  |  |
| --- | --- | --- | --- | --- | --- | --- | --- | --- | --- | --- | --- | --- | --- | --- |
|  | 15 - 24 | 25 - 34 | 35 - 44 | 45 - 54 | 55 - 64 | 65 - 74 | All | 15 - 24 | 25 - 34 | 35 - 44 | 45 - 54 | 55 - 64 | 65 - 74 | All |
| **Sex** |  |  |  |  |  |  |  |  |  |  |  |  |  |  |
| Male | 0.7 | 1.2 | 1.5 | 3.2 | 4.4 | 4.7 | 2.5 | 5.3 | 5.9 | 6.6 | 10.9 | 15.1 | 17.4 | 9.7 |
| Female | 0.6 | 0.8 | 2.2 | 2.4 | 3.9 | 4.7 | 2.3 | 5.4 | 5.5 | 8.2 | 10.1 | 14.5 | 16.9 | 9.8 |
| **Income Quintile** |  |  |  |  |  |  |  |  |  |  |  |  |  |  |
| Poorest | 0.9 | 2.5 | 5.3 | 9.3 | 12.3 | 8.5 | 5.8 | 6.8 | 11.2 | 15.5 | 24.9 | 33.5 | 28.3 | 18.6 |
| Q2 | 0.4 | 1.0 | 1.7 | 3.9 | 4.2 | 4.6 | 2.5 | 6.2 | 6.3 | 8.3 | 13.8 | 17.2 | 17.5 | 11.3 |
| Q3 | 0.6 | 1.0 | 1.6 | 1.9 | 3.6 | 3.3 | 2.0 | 4.5 | 5.4 | 6.2 | 9.3 | 13.8 | 14.5 | 8.6 |
| Q4 | 0.9 | 0.6 | 0.9 | 1.0 | 2.2 | 2.6 | 1.2 | 4.8 | 3.8 | 5.3 | 7.0 | 10.8 | 9.4 | 6.6 |
| Wealthiest | 0.5 | 0.2 | 0.7 | 0.8 | 1.4 | 2.3 | 0.9 | 3.4 | 2.5 | 4.0 | 5.0 | 6.3 | 8.8 | 4.7 |
| **Education** |  |  |  |  |  |  |  |  |  |  |  |  |  |  |
| Less than secondary school graduation | 2.1 | 4.2 | 8.8 | 7.7 | 9.7 | 6.8 | 7.1 | 13.5 | 20.0 | 21.6 | 27.5 | 29.7 | 29.9 | 26.2 |
| Post-secondary school graduation | 0.9 | 1.9 | 3.5 | 5.4 | 5.7 | 5.3 | 3.8 | 6.9 | 9.2 | 12.0 | 17.7 | 18.9 | 18.9 | 14.1 |
| More than post-secondary school | 0.6 | 0.8 | 1.4 | 2.1 | 3.3 | 4.1 | 1.9 | 4.7 | 4.7 | 6.4 | 8.4 | 12.5 | 14.0 | 7.9 |
| **Marital Status** |  |  |  |  |  |  |  |  |  |  |  |  |  |  |
| Married or common law | 0.8 | 0.8 | 1.3 | 2.1 | 3.4 | 4.2 | 2.2 | 7.9 | 4.3 | 5.7 | 8.5 | 12.8 | 15.6 | 9.0 |
| Other marital status | 0.7 | 1.3 | 3.6 | 4.9 | 6.2 | 5.9 | 2.7 | 5.1 | 7.6 | 12.7 | 16.4 | 20.4 | 20.9 | 10.9 |
| **Geography** |  |  |  |  |  |  |  |  |  |  |  |  |  |  |
| Urban | 0.7 | 1.0 | 1.9 | 2.7 | 4.1 | 4.9 | 2.4 | 5.5 | 5.8 | 7.4 | 10.4 | 14.9 | 17.5 | 9.7 |
| Rural | 0.5 | 0.9 | 1.8 | 3.0 | 4.2 | 3.8 | 2.6 | 4.4 | 4.9 | 7.3 | 10.9 | 14.2 | 16.0 | 10.0 |
| **Percent of Population Reporting Poor, Fair or Good Health** |  |  |  |  |  |  |  | **Percent of Population Reporting Poor, Fair, Good or Very Good Health** |  |  |  |  |  |  |
|  | 15 - 24 | 25 - 34 | 35 - 44 | 45 - 54 | 55 - 64 | 65 - 74 | All | 15 - 24 | 25 - 34 | 35 - 44 | 45 - 54 | 55 - 64 | 65 - 74 | All |
| **Sex** |  |  |  |  |  |  |  |  |  |  |  |  |  |  |
| Male | 29.1 | 31.2 | 33.6 | 39.7 | 45.7 | 50.1 | 37.5 | 70.1 | 71.7 | 75.1 | 79.6 | 81.6 | 82.9 | 76.5 |
| Female | 33.8 | 30.0 | 35.8 | 39.8 | 44.1 | 49.2 | 38.3 | 76.5 | 73.1 | 76.0 | 77.8 | 79.5 | 81.8 | 77.2 |
| **Income Quintile** |  |  |  |  |  |  |  |  |  |  |  |  |  |  |
| Poorest | 35.8 | 41.4 | 50.3 | 61.5 | 66.1 | 64.2 | 51.3 | 75.7 | 76.2 | 83.3 | 86.2 | 89.3 | 88.0 | 82.2 |
| Q2 | 35.4 | 36.7 | 39.1 | 47.8 | 52.4 | 51.7 | 43.5 | 75.8 | 75.7 | 77.5 | 81.2 | 82.9 | 84.1 | 79.4 |
| Q3 | 29.9 | 31.2 | 34.0 | 40.7 | 44.9 | 47.3 | 37.5 | 73.2 | 74.7 | 75.1 | 81.9 | 83.0 | 82.5 | 78.2 |
| Q4 | 27.0 | 24.6 | 30.3 | 34.5 | 39.5 | 40.5 | 32.2 | 71.8 | 70.9 | 73.5 | 77.3 | 78.8 | 79.4 | 75.0 |
| Wealthiest | 24.4 | 21.3 | 25.0 | 27.8 | 32.1 | 32.2 | 27.0 | 66.4 | 65.3 | 70.8 | 72.1 | 73.5 | 71.1 | 70.2 |
| **Education** |  |  |  |  |  |  |  |  |  |  |  |  |  |  |
| Less than secondary school graduation | 53.2 | 54.6 | 59.1 | 64.1 | 64.5 | 65.7 | 62.3 | 81.4 | 82.9 | 85.7 | 88.1 | 90.0 | 90.3 | 87.9 |
| Post-secondary school graduation | 37.8 | 40.9 | 43.5 | 50.5 | 52.0 | 54.0 | 46.6 | 79.4 | 78.0 | 80.4 | 85.1 | 84.5 | 85.3 | 82.3 |
| More than post-secondary school | 29.1 | 28.5 | 32.8 | 36.7 | 41.5 | 45.0 | 34.7 | 71.4 | 71.3 | 74.6 | 77.1 | 78.8 | 79.9 | 75.1 |
| **Marital Status** |  |  |  |  |  |  |  |  |  |  |  |  |  |  |
| Married or common law | 37.8 | 28.7 | 32.3 | 37.5 | 42.8 | 47.8 | 37.3 | 75.5 | 70.7 | 75.0 | 78.2 | 79.6 | 81.9 | 76.9 |
| Other marital status | 30.7 | 33.4 | 42.2 | 46.7 | 51.0 | 54.1 | 38.7 | 72.9 | 74.9 | 77.0 | 80.0 | 83.1 | 83.4 | 76.7 |
| **Geography** |  |  |  |  |  |  |  |  |  |  |  |  |  |  |
| Urban | 31.6 | 30.7 | 35.1 | 40.3 | 45.1 | 50.1 | 37.9 | 73.4 | 72.3 | 75.3 | 78.3 | 80.5 | 82.3 | 76.6 |
| Rural | 29.8 | 30.1 | 32.6 | 37.8 | 44.3 | 48.1 | 37.7 | 71.9 | 73.4 | 76.5 | 80.1 | 80.9 | 82.4 | 77.9 |

Notes: Darker values are greater values. Shading is independent within each dichotomization of self-reported health. Data Source: Pooled sample of the Canadian Community Health Survey 2013- 2015, Statistics Canada, author's calculations

Table 3 reports our regression results and our primary findings. The intercepts for all three models that we reported are high (between 0.92 and 0.96, p<0.001), suggesting that, on average, Canadians tend to report high levels of HUI. The unmediated effects of the sociodemographic variables suggest that HUI tends to be higher for younger and lower for older populations, lower for lower-income populations and higher for higher-income populations, lower for non-married or common law populations, lower for less educated populations, and lower for rural populations. Importantly, across all four estimated models, the dichotomization of self-rated health is associated with a negative change in HUI; however, this amount is greatest for the lowest cutpoint, “poor health” (−0.48, p<0.001), and least for the lowest cutpoint, “poor to very good health” (−0.06, p<0.001).

**Table 3.**
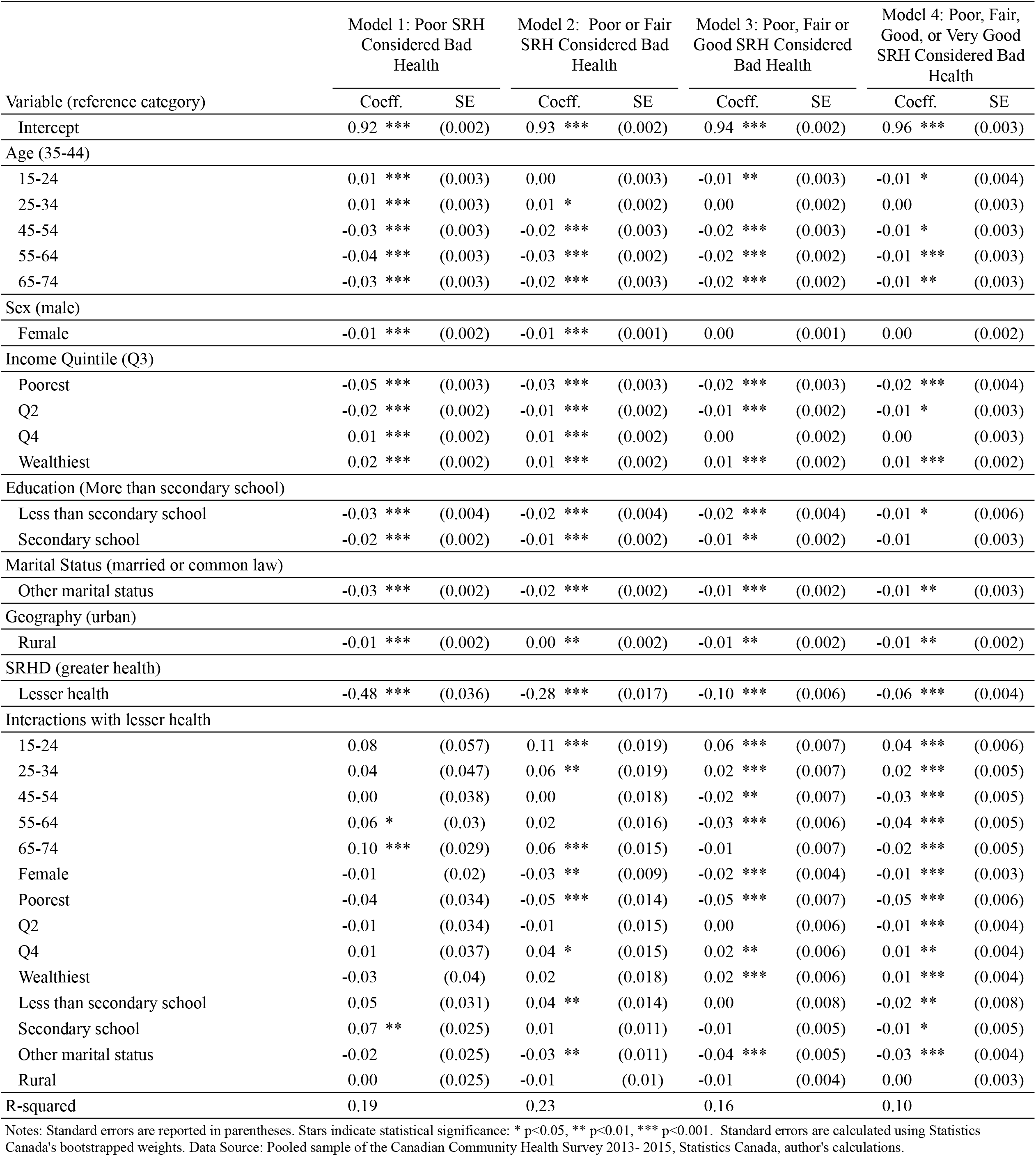
Regression results for different dichotomizations of self-rated health on the Health Utility Index in Canadian Community Health Survey, 2013-2015.

Interactions between the sociodemographic variables and self-rated health are more likely to have lower p values when the cutpoint used for dichotomization is higher and are consistently so for the highest cutpoint. However, the interaction effects of the oldest age groups, 55-64 (0.06, p<0.05) and 65-74 (0.10, p<0.001), and having only secondary education (0.07, p<0.01) have a p-value below 0.01, even for the lowest cutpoint. The direct effects of sociodemographic variables are also muted as the cutpoint is raised, further suggesting that their associations are mediated by it.

We were concerned that our findings might be patterned by large changes in the number of observations which fall on either side of the dichotomization of self-rated health as the cutpoint changes. To control for this, we also estimated our models using randomly selected samples of equal size on either side of each dichotomization and across samples. Sample sizes for these models were much smaller and had far fewer effects with p-values below 0.01. Nonetheless, the results were broadly similar. We provide these results in Appendix A.

## Discussion

Our results support two key findings that researchers should consider when dichotomizing self-rated health. First, in our representative sample of Canadians, dichotomizing self-rated health using lower cutpoints captured more pronounced differences in health-related quality of life, as measured by HUI. This suggests that working with lower cutpoints is likely to result in a more valid capture of discrepancies in health. In the extreme, as can be seen in Table 3, the distinction between “excellent” and “fair to very good health” captures virtually no difference in HUI at all (−0.06, p<0.001) after controlling for sociodemographic variables and their interactions with this distinction. Second, using lower cutpoints also tended to capture more consistent differences in HUI status across subpopulations; however, reliability remained an issue, particularly for older populations. This result is consistent with the finding that older populations tend to underweight their health when they self-rate.^3,23^ In summary, dichotomizing SRH using lower cut points results in differences in SRH being more likely to reflect differences in levels of HUI between populations rather than systematic differences in how respondents tend to interpret the SRH question. Said differently, our findings suggest that diverse groups of people tend to agree on what poor health looks like but are less likely to agree on what good health looks like.

Overall, our findings suggest that using lower cutpoints when dichotomizing self-reported health results in more valid and reliable estimates. However, using lower cutpoints also means working with health outcomes that are far less prevalent and, therefore, more difficult to estimate for smaller populations or to estimate accurately in smaller samples. This likely explains the common practice in the literature of using much higher cutpoints. Our results should caution researchers against doing so blindly.

### Limitations

With its inherent limitations and focus on function, we are mindful that HUI is only one measure of health that could be used to validate self-rated health dichotomization. Therefore, the strength of our conclusions could greatly benefit from future replication using different measures of health status and different populations, ideally using measures that are not also self-reported, like the HUI. Additionally, we only consider one measure of a Likert scale of self-rated health. To take just one example, in their Finish study, Finnäs et al.^14^ work with a Likert scale that ranges between “very poor,” “fairly poor,” “average,” “fairly good,” and “good.” It is unclear how this scale maps to the Canadian scale; for example, are the middle responses “fair” and “average” equivalent? Although the patterns we observe in our results appear incremental as we move from one end of the Likert scale to the other, it is unclear whether this pattern will be the same for every articulation of self-rated health.^24,25^

## Conclusion

Our study provides an important corrective against blindly applying dichotomizations to self-reported health in population health research and reporting. Nonetheless, the strength of our conclusion could greatly benefit from future replication in additional populations and using different health status measures. Self-reported health indicators are only sometimes collected alongside other leading non-self-reported indicators in administrative and registry data, such as hospitalization rates or life expectancy. However, new data linkages by Statistics Canada between the CCHS and these administrative and registry sources promise to support the replication and extension of our analysis.

## Author Contributions

CP & SM contributed to the study conception, methods, analysis, and writing. CN contributed to the study conception and discussion. All authors provided critical feedback and helped shape the research, analysis and manuscript editing. All authors approved the final version of the manuscript.

## Acknowledgments

This research was conducted at The Saskatchewan Research Data Centre, which is part of the Canadian Research Data Centre Network (CRDCN). This service is provided through the support of the University of Saskatchewan, the Province of Saskatchewan, the Canadian Foundation for Innovation, the Canadian Institutes of Health Research, the Social Science and Humanity Research Council, and Statistics Canada. All views expressed in this work are our own. The authors would also like to thank our RDC analyst Ruben Mercado, especially for his help and support and Dr. Grant Gibson for his insight and comments. Finally, we thank our anonymous reviewers for their helpful suggestions.

## Data Availability

All data used in this study is available through Statistics Canada’s Research Data Centres program.

---

**Appendix A.** Regression results for different dichotomizations of self-rated health on the Health Utility Index, for randomly selected groups of equal size on either side of dichotimizations of self-rated health in Canadian Community Health Survey, 2013-2015.

| Variable (reference category) | Model 1: Poor SRH<br>Considered Bad<br>Health |  | Model 2: Poor or Fair<br>SRH Considered Bad<br>Health |  | Model 3: Poor, Fair or<br>Good SRH Considered<br>Bad Health |  | Model 4: Poor, Fair,<br>Good, or Very Good<br>SRH Considered Bad<br>Health |  |
| --- | --- | --- | --- | --- | --- | --- | --- | --- |
|  | Coeff. | SE | Coeff. | SE | Coeff. | SE | Coeff. | SE |
| Intercept | 0.93 *** | (0.012) | 0.94 *** | (0.011) | 0.95 *** | (0.007) | 0.96 *** | (0.008) |
| Age (35-44) |  |  |  |  |  |  |  |  |
| 15-24 | 0.02 | (0.015) | -0.01 | (0.014) | 0.00 | (0.012) | -0.02 | (0.015) |
| 25-34 | 0.02 | (0.012) | 0.02 | (0.009) | 0.00 | (0.008) | 0.00 | (0.008) |
| 45-54 | -0.03 * | (0.015) | -0.03 * | (0.015) | -0.02 * | (0.012) |  | (0) |
| 55-64 | -0.04 ** | (0.013) | -0.05 *** | (0.014) | -0.02 * | (0.007) | -0.01 | (0.009) |
| 65-74 | -0.02 | (0.015) | -0.02 | (0.013) | -0.02 * | (0.008) | -0.01 | (0.007) |
| Sex (male) |  |  |  |  |  |  |  |  |
| Female | -0.02 * | (0.008) | -0.01 | (0.008) | 0.00 | (0.007) | -0.01 | (0.006) |
| Income Quintile (Q3) |  |  |  |  |  |  |  |  |
| Poorest | -0.06 *** | (0.015) | -0.03 * | (0.013) | -0.02 | (0.011) | -0.02 | (0.014) |
| Q2 | 0.00 | (0.013) | -0.04 * | (0.015) | 0.00 | (0.008) | 0.00 | (0.007) |
| Q4 | 0.01 | (0.012) | 0.01 | (0.009) | 0.00 | (0.009) | 0.00 | (0.008) |
| Wealthiest | 0.03 * | (0.011) | 0.01 | (0.009) | 0.00 | (0.011) | 0.01 | (0.006) |
| Education (More than secondary school) |  |  |  |  |  |  |  |  |
| Less than secondary school | -0.03 | (0.023) | -0.05 | (0.03) | -0.01 | (0.014) | -0.02 | (0.016) |
| Secondary school | -0.02 | (0.012) | -0.02 | (0.011) | 0.00 | (0.008) | -0.02 | (0.011) |
| Marital Status (married or common law) |  |  |  |  |  |  |  |  |
| Other marital status | -0.03 ** | (0.01) | -0.01 | (0.01) | -0.03 *** | (0.008) | 0.00 | (0.008) |
| Geography (urban) |  |  |  |  |  |  |  |  |
| Rural | -0.03 ** | (0.011) | 0.00 | (0.009) | 0.00 | (0.007) | 0.00 | (0.005) |
| SRHD (greater health) |  |  |  |  |  |  |  |  |
| Lesser health | -0.49 *** | (0.038) | -0.29 *** | (0.039) | -0.12 *** | (0.023) | -0.06 *** | (0.017) |
| Interactions with lesser health |  |  |  |  |  |  |  |  |
| 15-24 | 0.08 | (0.058) | 0.13 ** | (0.044) | 0.09 *** | (0.024) | 0.08 ** | (0.029) |
| 25-34 | 0.03 | (0.047) | 0.01 | (0.041) | 0.01 | (0.026) | 0.03 | (0.02) |
| 45-54 | 0.01 | (0.041) | -0.01 | (0.038) | 0.02 | (0.025) | -0.01 | (0.018) |
| 55-64 | 0.07 * | (0.032) | 0.05 | (0.033) | -0.03 | (0.021) | -0.04 * | (0.02) |
| 65-74 | 0.09 ** | (0.032) | 0.04 | (0.035) | 0.01 | (0.021) | -0.03 | (0.02) |
| Female | 0.00 | (0.022) | -0.03 | (0.019) | -0.02 | (0.014) | -0.01 | (0.011) |
| Poorest | -0.03 | (0.038) | -0.02 | (0.028) | -0.02 | (0.022) | -0.08 ** | (0.025) |
| Q2 | -0.03 | (0.038) | 0.02 | (0.031) | 0.00 | (0.022) | -0.01 | (0.017) |
| Q4 | 0.01 | (0.039) | 0.04 | (0.029) | 0.04 | (0.022) | 0.02 | (0.015) |
| Wealthiest | -0.03 | (0.042) | 0.02 | (0.033) | 0.02 | (0.025) | 0.01 | (0.015) |
| Less than secondary school | 0.05 | (0.036) | 0.04 | (0.039) | -0.03 | (0.029) | 0.01 | (0.027) |
| Secondary school | 0.07 * | (0.027) | 0.03 | (0.022) | -0.02 | (0.02) | 0.01 | (0.018) |
| Other marital status | -0.02 | (0.026) | -0.01 | (0.022) | -0.03 | (0.017) | -0.04 * | (0.018) |
| Rural | 0.02 | (0.026) | -0.01 | (0.019) | -0.02 | (0.019) | -0.01 | (0.011) |
| R-squared | 0.49 |  | 0.30 |  | 0.16 |  | 0.14 |  |

Notes: Standard errors are reported in parentheses. Stars indicate statistical significance: \* p<0.05, \*\* p<0.01, \*\*\* p<0.001. Standard errors are calculated using Statistics Canada's bootstrapped weights. Data Source: Pooled sample of the Canadian Community Health Survey 2013- 2015, Statistics Canada, author's calculations.

## Footnotes

1 van Doorslaer and Jones^10^ posit HUI as a compelling measure of “‘actual’ health” and use it to explore the functional form of self-rated health in the Canadian National Population Health Survey (a precursor to the CCHS).

2 Together, these exclusions represent about 3% of the Canadian population.

## References

1. Wu, S. et al. The relationship between self-rated health and objective health status: a population-based study. BMC Public Health 13, 320 (2013).

2. Schnittker, J. & Bacak, V. The increasing predictive validity of self-rated health. PLoS One 9, e84933 (2014).

3. Falconer, J. & Quesnel-Vallée, A. The moderating effect of sociodemographic factors on the predictive power of self-rated health for mortality in Canada. Can. Stud. Popul. 44, 77–99 (2017).

4. Vaillant, N. & Wolff, F.-C. On the reliability of self-reported health: evidence from Albanian data. J. Epidemiol. Glob. Health 2, 83–98 (2012).

5. Miilunpalo, S., Vuori, I., Oja, P., Pasanen, M. & Urponen, H. Self-rated Health Status as a Health Measure: The Predictive Value of Self-Reported Health Status on the Use of Physician Services and on Mortality in the Working-Age Population. J. Clin. Epidemiol. 50, 517–528 (1997).

6. Burström, B. & Fredlund, P. Self rated health: Is it as good a predictor of subsequent mortality among adults in lower as well as in higher social classes? J. Epidemiol. Community Health 55, 836–840 (2001).

7. Zajacova, A. & Dowd, J. B. Reliability of Self-rated Health in US Adults. American Journal of Epidemiology vol. 174 977–983 Preprint at 10.1093/aje/kwr204 (2011).

8. Kananen, L. et al. Self-rated health in individuals with and without disease is associated with multiple biomarkers representing multiple biological domains. Sci. Rep. 11, 6139 (2021).

9. Jylhä, M. What is self-rated health and why does it predict mortality? Towards a unified conceptual model. Soc. Sci. Med. 69, 307–316 (2009).

10. van Doorslaer, E. & Jones, A. M. Inequalities in self-reported health: validation of a new approach to measurement. J. Health Econ. 22, 61–87 (2003).

11. CIHI. Reducing Gaps in Health: A Focus on Socio-Economic Status in Urban Canada. (2008).

12. Horsman, J., Furlong, W., Feeny, D. & Torrance, G. The Health Utilities Index (HUI): concepts, measurement properties and applications. Health Qual. Life Outcomes 1, 54 (2003).

13. Furlong, W. J. & McMaster University. Centre for Health Economics and Policy Analysis. The Health Utilities Index (HUI) System for Assessing Health-Related Quality of Life in Clinical Studies [electronic Resource]. (Hamilton, Ont.: Centre for Health Economics and Policy Analysis, McMaster University, 2001).

14. Finnas, F., Nyqvist, F. & Saarela, J. Some methodological remarks on self-rated health. Open Public Health J. 1, 32–39 (2009).

15. Bourne, P. A. Dichotomising poor self-reported health status: Using secondary cross-sectional survey data for Jamaica. N. Am. J. Med. Sci. 1, 295–302 (2009).

16. Sung, L. et al. Construct validation of the Health Utilities Index and the Child Health Questionnaire in children undergoing cancer chemotherapy. Br. J. Cancer 88, 1185–1190 (2003).

17. Davison, S. N., Jhangri, G. S. & Feeny, D. H. Comparing the Health Utilities Index Mark 3 (HUI3) with the Short Form-36 preference-based SF-6D in chronic kidney disease. Value Health 12, 340–345 (2009).

18. Feng, Y., Bernier, J., McIntosh, C. & Orpana, H. Validation of disability categories derived from Health Utilities Index Mark 3 scores. Health Rep. 20, 43–50 (2009).

19. Feeny, D., Kaplan, M. S., Huguet, N. & McFarland, B. H. Comparing population health in the United States and Canada. Popul. Health Metr. 8, 8 (2010).

20. Furlong, W. J., Feeny, D. H., Torrance, G. W. & Barr, R. D. The Health Utilities Index (HUI®) system for assessing health-related quality of life in clinical studies. Ann. Med. 33, 375–384 (2001).

21. Kopec, J. A., Schultz, S. E., Goel, V. & Ivan Williams, J. Can the health utilities index measure change? Med. Care 39, 562–574 (2001).

22. Torrance, G. & Feeny, D. The Health Utilities Index. (1990).

23. Zajacova, A. & Woo, H. Examination of Age Variations in the Predictive Validity of Self-Rated Health. J. Gerontol. B Psychol. Sci. Soc. Sci. 71, 551–557 (2016).

24. Eriksson, I., Undén, A. L. & Elofsson, S. Self-rated health. Comparisons between three different measures. Results from a population study. Int. J. Epidemiol. 30, 326–333 (2001).

25. Cullati, S. et al. Does the single-item self-rated health measure the same thing across different wordings? Construct validity study. Qual. Life Res. 29, 2593–2604 (2020).

